# Natural course of post COVID-19 condition and implications for trial design and outcome selection: A population-based longitudinal cohort study

**DOI:** 10.1101/2022.06.22.22276746

**Authors:** Tala Ballouz, Dominik Menges, Alexia Anagnostopoulos, Anja Domenghino, Hélène E Aschmann, Anja Frei, Jan S Fehr, Milo A Puhan

**Author notes:** **Registration**: ISRCTN14990068. **Funding source**: Swiss Federal Office of Public Health, Department of Health of the Canton of Zurich, University of Zurich Foundation.

## Abstract

**Background:** Evidence from population-based studies on the longer-term natural course of post COVID-19 condition is limited, but crucial for informing patients and healthcare providers and for effectively designing clinical trials.

**Objectives:** To evaluate longer-term symptoms and health outcomes within a cohort of SARS-CoV-2 infected individuals.

**Design:** Population-based, longitudinal cohort.

**Setting:** General population, Canton of Zurich, Switzerland.

**Patients:** 1543 adults with confirmed SARS-CoV-2 infection and 628 adults without infection.

**Measurements:** Changes in self-reported health status over time, factors associated with persistence of non-recovery, and prevalence and excess risk of symptoms at 6 and 12 months post-infection compared to non-infected individuals.

**Results:** 25% of SARS-CoV-2 infected individuals did not recover by 6 months. Of those, 67% and 58% also did not recover at 12 and 18 months after infection, respectively. Hospitalization for acute COVID-19, pre-existing fatigue and pain or discomfort, and presence of specific systemic, cardiovascular, or musculoskeletal symptoms at 6 months were associated with persistent non-recovery. Symptom prevalence was higher among infected individuals compared to non-infected individuals at 6 months (adjusted risk difference (aRD)=17%) and 12 months (aRD=20%). aRDs for individual symptoms ranged from 2% to 12%, with the highest excess risks observed for altered taste or smell, post-exertional malaise, fatigue, and reduced concentration and memory.

**Limitations:** We relied on self-reported assessments and did not assess the effects of vaccination or infection with emerging variants of concern.

**Conclusion:** These findings emphasize the need for effective interventions to reduce the burden of post COVID-19 condition. They further demonstrate the importance of using multiple outcome measures and of considering the expected rates of natural recovery and heterogenous patient trajectories in the design and interpretation of clinical trials.

## Introduction

Post COVID-19 condition affects approximately 20-30% of unvaccinated individuals at three to six months after SARS-CoV-2 infection (1–10). To adequately inform patients, healthcare providers and policy makers, it is important to not only determine its prevalence but also its natural course over time (11). As evidence on the substantial public health burden of post COVID-19 condition accumulates, clinical trials will be necessary to establish interventions which accelerate recovery or provide relief for associated symptoms. A solid understanding of post COVID-19 condition trajectories and relevant outcome measures is required to effectively design and adequately interpret such trials.

To date, few studies have reported outcomes related to post COVID-19 condition beyond six months after infection (3,12–16). Common study limitations include specific participant populations (e.g., hospitalized) or the lack of a prospective follow-up. Thus, these studies may not adequately reflect the variability of symptoms and recovery across the severity spectrum of acute COVID-19. Additionally, most studies lacked a comparison with a comparator group, and their findings were often scrutinized as many reported symptoms were non-specific and common in the general population without infection. Furthermore, limited knowledge and the lack of consensus on core outcomes regarding post COVID-19 condition has led to the use of various outcome measures and inconsistent symptom definitions across observational studies affecting their comparability (17). This also impairs the interpretation of future trials of interventions aiming to improve post COVID-19 condition and their translation into healthcare policy and clinical practice.

In this study, we aimed to characterize the natural course of post COVID-19 condition within a population-based, longitudinal cohort of SARS-CoV-2 infected individuals. Specifically, our objectives were to 1) describe patterns of recovery and symptom persistence over 12-18 months, 2) determine the attributable risk of related symptoms by comparing their prevalence in infected individuals and a general population cohort with no evidence of a past infection, and 3) evaluate factors potentially associated with non-recovery. Thereby, we aim to provide comprehensive evidence on different trajectories and relevant outcomes of post COVID-19 condition to support the design and interpretation of future clinical trials.

## Methods

A detailed description of the methods is provided in the Supplementary Material.

### Study design and populations

This analysis is based on the Zurich SARS-CoV-2 Cohort, an ongoing, population-based, prospective study of individuals with laboratory-confirmed SARS-CoV-2 infection, recruited through the Department of Health of the Canton of Zurich. All participants provided written consent. The study was prospectively registered (ISRCTN14990068) and approved by the ethics committee of the Canton of Zurich (BASEC 2020-01739).

We enrolled two populations of SARS-CoV-2 infected individuals: 1) “*retrospectively recruited”* which included individuals infected with SARS-CoV-2 prior to study initiation (27 February to 05 August 2020) and enrolled at a median of 7.2 months after infection, and 2) “*prospectively recruited”* which included an age-stratified random sample of individuals diagnosed between 06 August 2020 and 19 January 2021, enrolled upon diagnosis. All participants were enrolled before the roll-out of COVID-19 vaccines and emergence of variants of concern in Switzerland.

To compare health outcomes among SARS-CoV-2 infected individuals with those in a similar uninfected control population, we leveraged data from phase IV of the Corona Immunitas Zurich seroprevalence study ((18,19); ISRCTN18181860, BASEC 2020-01247) which consists of an age-stratified random sample of individuals from the general population of the Canton of Zurich. We excluded participants who were infected in the past or at the time of enrolment.

### Data sources

We used data collected in the Zurich SARS-CoV-2 Cohort upon enrolment and at regularly conducted follow-ups up to 12 months after infection. Initially, participants consented to participation for one year and were asked to consent for further biannual assessments upon completion of the 12-month assessment. At the time of writing, complete 18-month data was available only for retrospectively recruited participants (77% agreed to prolongation and completed 18-month questionnaire). To provide a longer perspective of the course of post COVID-19 condition, we also included 18-month outcomes from these participants.

Furthermore, we included data from the baseline questionnaire of the Corona Immunitas study, which was closely aligned with the Zurich SARS-CoV-2 Cohort questionnaires to ensure comparability. The timeframe of data collection in Corona Immunitas (May-August 2021) corresponded to the timeframe of the 6-month and 12-month assessments of the prospectively and retrospectively recruited populations in the Zurich SARS-CoV-2 Cohort, respectively.

### Main outcomes

In the absence of an established core outcome set for post COVID-19 condition, we determined outcomes based on commonly reported patient-relevant health measures and symptoms (17,20).

Our primary outcome was the overall relative health status of participants at 6 and 12 months after infection, defined using a combination of two self-reported measurements: 1) *(non-)recovery status* compared to usual health status before infection, and 2) *overall health* using the EuroQol visual analogue scale (EQ-VAS), with cut-off values for different levels of health impairment determined based on population normative values and studies on chronic-obstructive pulmonary disease (21–23).

Secondary outcomes included change in relative health status between the 6- and 12-month follow-up and the presence and severity of 23 predefined symptoms at both follow-up timepoints. We also asked participants to indicate whether they perceived symptoms to be related to COVID-19 to differentiate post-COVID-19 symptoms from those due to other reasons. Additional scale-based assessments included fatigue (Fatigue Assessment Scale [FAS]), dyspnea (modified Medical Research Council [mMRC] dyspnea scale), depression, anxiety, and stress (21-item Depression, Anxiety, and Stress Scale [DASS-21]), and health-related quality of life (HRQoL; EuroQol 5-dimension 5-level [EQ-5D-5L]). Calculation of scores for these assessments followed official guidance, as described previously (5,24).

### Statistical analysis

In line with our objectives, we conducted three main analyses. First, we descriptively analyzed the overall relative health status among infected individuals (all Zurich SARS-CoV-2 Cohort participants) at the 6-month follow-up and determined the proportion with complete recovery or mild, moderate, and severe health impairment. We performed multivariable regression analyses to assess the influence of potential predictors on non-recovery by 6 months. Furthermore, we calculated the prevalence with 95% confidence interval (CI) for each of the symptoms at 6 and 12 months, grouped based on the affected organ system. Second, we estimated the excess risk of symptoms and adverse health outcomes (adjusted risk difference [aRD]) in infected individuals (Zurich SARS-CoV-2 Cohort) 6 and 12 months after infection compared to the general population (Corona Immunitas Zurich) using logistic regression models. Since the data collection timeframe of Corona Immunitas corresponded to different follow-up timepoints of the prospectively (6-month) and retrospectively recruited (12-month) participants, we performed these comparative analyses separately for the two groups to minimize the influence of the timing of assessments on measured outcomes.

Third, we focused on longer-term outcome trajectories among SARS-CoV-2 infected participants within the Zurich SARS-CoV-2 Cohort who reported non-recovery at 6 months after infection. We evaluated the change in relative health status at 12 months and assessed associations of potential predictors for persistent non-recovery using logistic regression models. We descriptively analyzed the proportions of participants reporting symptoms at the 6- and 12-month follow-ups and evaluated their co-occurrence. Further, we compared the proportion of participants with adverse health outcomes prior to COVID-19 and up to 12 months in non-recovered participants to those recovering by 6 months. Finally, we descriptively analyzed the longitudinal trajectories of the aforementioned outcomes at 18 months among retrospectively recruited participants reporting non-recovery at 6 months.

### Role of the funding source

This study is part of Corona Immunitas research network, coordinated by the Swiss School of Public Health (SSPH+), and funded by fundraising of SSPH+ including funds of the Swiss Federal Office of Public Health and private funders (ethical guidelines for funding stated by SSPH+ were respected), by funds of the Cantons of Switzerland (Vaud, Zurich, and Basel) and by institutional funds of the Universities. Additional funding specific to this study was received from the Department of Health of the Canton of Zurich, the University of Zurich Foundation, and the Swiss Federal Office of Public Health. The funding bodies had no influence on the design, conduct, analysis, or interpretation of the study, as well as on the decision to publish, preparation or revisions of the manuscript. TB received funding from the European Union’s Horizon 2020 research and innovation programme under the Marie Skłodowska-Curie grant agreement No 801076, through the SSPH+ Global PhD Fellowship Programme in Public Health Sciences (GlobalP3HS) of the SSPH+. HA received a Swiss National Science Foundation (SNSF) Early Postdoc. Mobility Fellowship.

## Results

We included data from 1543 SARS-CoV-2 infected individuals (Zurich SARS-CoV-2 Cohort) and 628 non-infected individuals (Corona Immunitas) (Table 1; Supplementary Table 1 & Figure 1). Participants enrolled in the Zurich SARS-CoV-2 Cohort were on average younger than those in Corona Immunitas (49 versus 65 years). 51% of all participants were female.

**Table 1.** Characteristics of participants enrolled in the Zurich SARS-CoV-2 Cohort study (infected individuals) and phase IV of the Corona Immunitas Zurich seroprevalence study (control population).

|  | SARS-CoV-2 infected individuals<br>(Zurich SARS-CoV-2 Cohort)<br>(N=1543) | Non-infected individuals in control<br>population<br>(Corona Immunitas Zurich)<br>(N=628) |
| --- | --- | --- |
| **Age (years), Median (IQR)** | 49.0 (34.0 to 63.0) | 65.0 (45.0 to 72.0) |
| **Age group** |  |  |
| 18-39 years | 510 (33.1%) | 128 (20.4%) |
| 40-64 years | 656 (42.5%) | 174 (27.7%) |
| 65+ years | 377 (24.4%) | 326 (51.9%) |
| **Female sex** | 781 (50.6%) | 322 (51.3%) |
| **Initial symptom severity** |  |  |
| Asymptomatic | 195 (12.8%) | - |
| Mild to moderate | 942 (61.8%) | - |
| Severe to very severe | 388 (25.4%) | - |
| Missing | 18 | - |
| **Initial symptom count, Median (IQR)** | 5.0 (3.0 to 8.0) | - |
| **Hospitalization and ICU stay** |  |  |
| Non-hospitalized | 1401 (91.5%) | - |
| Hospitalized without ICU stay | 115 (7.5%) | - |
| Hospitalized with ICU stay | 15 (1.0%) | - |
| Intubation during ICU stay | 8 (53.3%) | - |
| Missing | 12 | - |
| **At least one medical comorbidity** | 471 (30.8%) | 225 (35.8%) |
| Missing | 16 | 0 |
| **Monthly household income** |  |  |
| <6000 CHF | 489 (33.8%) | 219 (38.1%) |
| 6000 - 12000 CHF | 614 (42.4%) | 234 (40.8%) |
| >12000 CHF | 344 (23.8%) | 121 (21.1%) |
| Missing | 96 | 55 |
| **Current employment status** |  |  |
| Employed | 1027 (67.7%) | 255 (40.7%) |
| Student | 67 (4.4%) | 25 (4.1%) |
| Retired | 327 (21.5%) | 322 (51.4%) |
| Unemployed or other | 97 (6.4%) | 24 (3.8%) |
| Missing | 25 | 2 |
| **Highest education level reached** |  |  |
| None or mandatory school | 65 (4.3%) | 32 (5.1%) |
| Vocational training or specialized<br>baccalaureate | 645 (42.5%) | 281 (45.0%) |
| Higher technical school or<br>college | 395 (26.0%) | 159 (25.5%) |
| University | 412 (27.2%) | 152 (24.4%) |
| Missing | 26 | 4 |
| **Swiss nationality** | 1309 (84.8%) | 530 (84.4%) |
| **Smoking status** |  |  |
| Non-smoker | 901 (59.3%) | 336 (53.6%) |
| Ex-smoker | 410 (27.0%) | 208 (33.2%) |
| Smoker | 209 (13.8%) | 83 (13.2%) |
| Missing | 23 | 1 |
| **Body mass index, Median (IQR)** | 24.4 (22.0 to 26.8) | 24.2 (21.9 to 26.9) |

IQR: interquartile range; ICU: intensive care unit; CHF: Swiss francs

In total, 87% (1336/1543) of infected individuals were symptomatic and 8% (130/1543) were hospitalized during acute infection. Compared to prospectively recruited participants (4%), a higher proportion of retrospectively recruited (19%) were hospitalized, likely due to restriction of SARS-CoV-2 testing at the beginning of the pandemic to high-risk and symptomatic individuals.

### Non-recovery and symptoms among SARS-CoV-2 infected

Overall, 49% (699/1416, 127 with missing data) reported that they returned to their normal health status in less than a month after infection, while 18% (250/1416) reported recovery within 1 to 3 months. By 6 months, 25% (348/1418, 125 with missing data) of participants reported that they had not yet recovered with 17%, 4% and 3% with mild, moderate, and severe health impairment, respectively (1% missing EQ-VAS data). In multivariable regression models, older age, female sex, presence of comorbidities, and increased symptom count during acute infection were associated with non-recovery at 6 months (Supplementary Table 4 & Figure 2).

At the 6- and 12-month follow-ups, 47% (95% CI: 45% to 50%) and 50% (47% to 52%) of participants, respectively, reported experiencing any of the symptoms (Figure 1, Supplementary Figure 3). Meanwhile, the prevalence of any symptom considered to be related to COVID-19 was 27% (25% to 30%) at 6 months and 25% (23% to 28%) at 12 months. At both follow-ups, systemic (specifically fatigue, post-exertional malaise), respiratory (dyspnea), neurologic (headache, altered taste or smell, sleep disturbances, concentration difficulties), and musculoskeletal (myalgia) symptoms were most frequently reported with a decreased prevalence at 12 months. Self-reported symptom severity for most symptoms decreased or remained unchanged over time.

**Figure 1.**
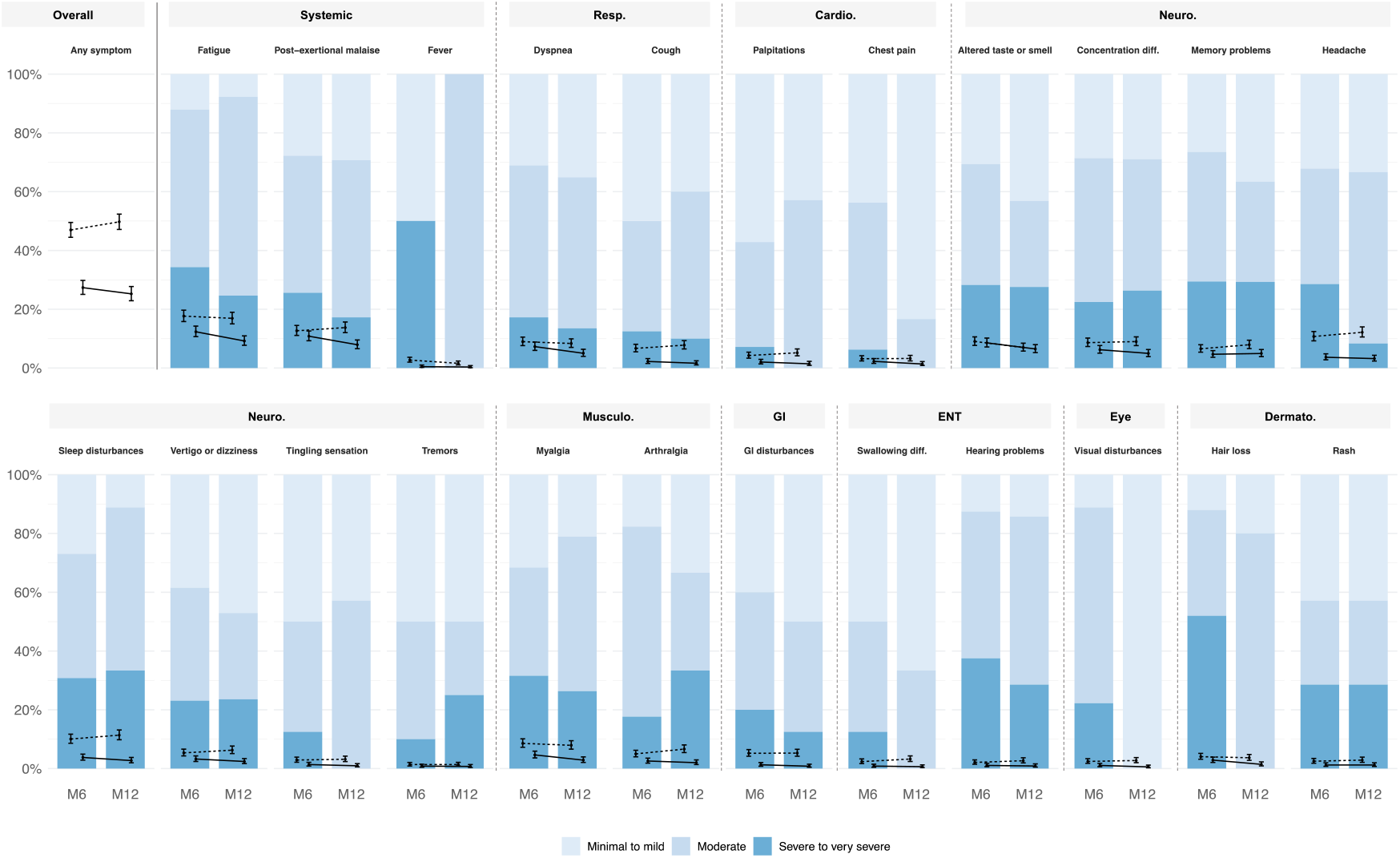
Prevalence and severity of 23 prespecified symptoms at 6 and 12 months after SARS-CoV-2 infection, grouped by body organ system affected. Error bars show the prevalence and 95% confidence interval for each of the symptoms among all participants in the Zurich SARS-CoV-2 Cohort (N=1543). Solid line refers to symptoms reported by the participants to be related to COVID-19. Dotted line refers to symptoms regardless of their relation to COVID-19. Stacked bar plot shows self-reported severity of symptoms in the prospectively recruited cohort (N=1106). M6: 6 months after infection, M12: 12 months, Resp.: respiratory, Cardio.: cardiovascular, Neuro.: neurologic, Musculo.: musculoskeletal, GI: gastrointestinal, ENT: ear, nose and throat, Dermato: dermatologic, diff.: difficulties.

### Symptoms attributable to infection

When comparing symptom prevalence among SARS-CoV-2 infected individuals with non-infected individuals from the general population, we found evidence of higher symptom prevalence among those infected at 6 months (aRD=17%, 11% to 21%) and 12 months (aRD=20%, 14% to 26%) after infection. The highest excess risks among infected individuals were for altered taste or smell, post-exertional malaise, fatigue, reduced concentration or memory, dyspnea, palpitations, chest pain, hair loss, sleep disturbances, and swallowing difficulties, with aRDs ranging between 2% and 12% (Figure 2, Supplementary Table 5).

**Figure 2.**
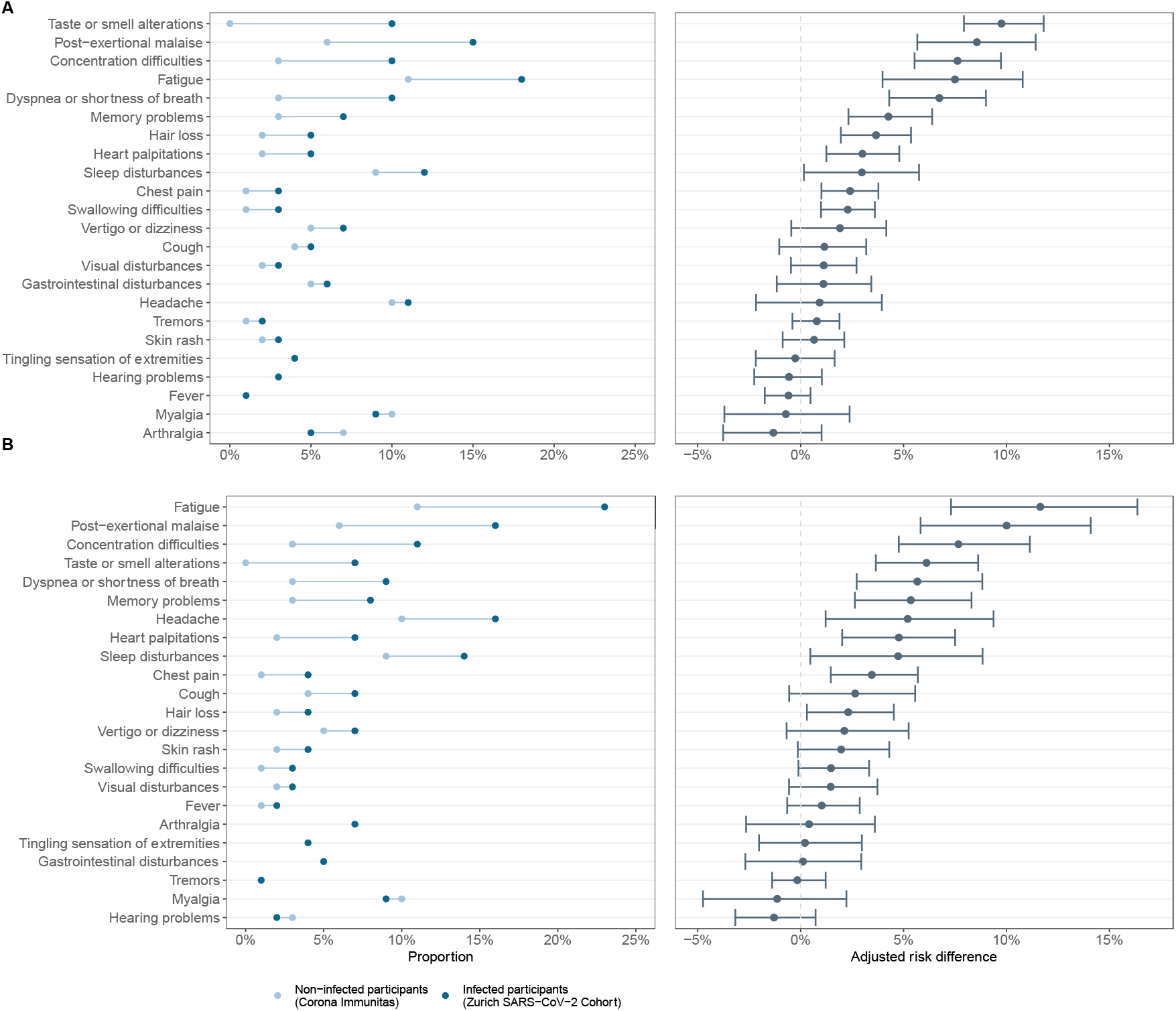
(A) Observed proportions and excess risk (adjusted risk differences) of each of the symptoms at 6 months after infection in the prospectively recruited infected group (N=1106; Zurich SARS-CoV-2 Cohort) compared to non-infected individuals from the general population (N=628; Corona Immunitas Zurich), as estimated based on logistic regression models adjusted for age groups, sex, comorbidity and body mass index. (B) Observed proportions and excess risk of each of the symptoms at 12 months after infection in the retrospectively recruited infected group (N=437; Zurich SARS-CoV-2 Cohort) compared to non-infected individuals (N=628; Corona Immunitas Zurich).

When evaluating adverse health outcomes (Supplementary Figure 4 & Table 5), we found strong evidence for a higher proportion of participants experiencing symptoms of anxiety among infected individuals at 6 months (aRD=5%, 2% to 8%) and 12 months (aRD=12%, 8% to 16%) based on DASS-21. We also found evidence that a higher proportion of infected individuals experienced anxiety or depression (aRD=7%, 2% to 13%) and problems during usual activities (aRD=8%, 4% to 12%) at 12 months based on EQ-5D-5L. Meanwhile, a higher proportion of non-infected individuals experienced pain or discomfort at both timepoints (6-month aRD=11%, 6% to 15%; 12-month aRD=9%, 3% to 15%).

### Symptoms and health trajectories among non-recovered

After 12 months, 63% (57% to 68%) of those reporting non-recovery at 6 months (i.e., 16% overall) reported not having fully recovered. However, there was a decrease in the proportion of those with mild, moderate, and severe health impairment to 11%, 3%, and 1%, respectively (1% missing EQ-VAS data). When assessing changes in the overall relative health status, most participants (53%) improved, particularly among those experiencing a moderate or severe health impairment at 6 months (Supplementary Figure 5). Yet, the health status of some individuals did not change (36%) and even worsened for a smaller proportion (11%).

Among non-recovered, fatigue (38% at 6-month and 25% at 12-month follow-up), post-exertional malaise (33% and 23%), altered taste or smell (23% and 16%), dyspnea (22% and 14%), reduced concentration (19% and 15%) or memory (13% and 14%), and myalgia (13% and 7%) were the most commonly reported COVID-19 related symptoms. Symptoms mostly decreased in severity over time (Supplementary Figure 6). Although most participants reporting COVID-19 related symptoms also reported non-recovery at 12 months, 11% nevertheless stated that they had fully recovered. Among those with at least two body organ systems involved (Supplementary Figure 7), systemic and neurologic symptoms co-occurred most frequently, followed by combinations of systemic, neurologic, and respiratory symptoms.

Longitudinal assessment of adverse health outcomes compared to pre-COVID-19 levels was available for prospectively recruited participants (Figure 3). Compared to those recovering by 6 months, higher proportions of non-recovered participants reported adverse health outcomes at all timepoints, including prior to infection. In both groups, there was an increase in the proportion of participants reporting adverse health outcomes shortly after infection. While this proportion decreased and returned to pre-COVID-19 levels among those who recovered, there was a slower decrease among those who did not recover. At 12 months, all outcomes were still reported by a considerable proportion of participants (57% with fatigue on FAS, 43% with dyspnea grade ≥1 on mMRC, 24% with depression, 27% with anxiety and 20% with stress on DASS-21, and 66% with problems on EQ-5D-5L).

**Figure 3.**
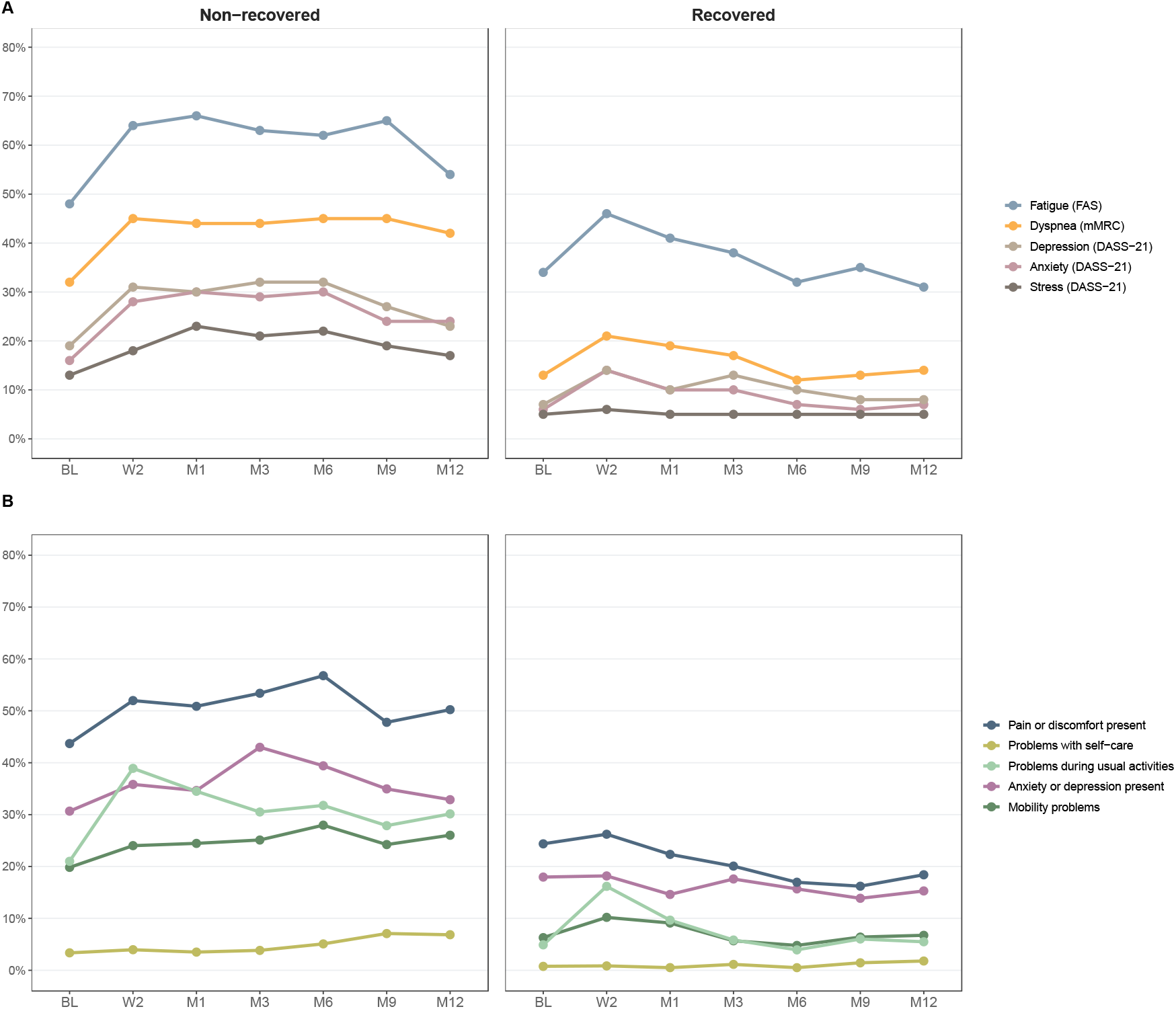
Adverse health outcomes stratified by recovery status at 6 months after SARS-CoV-2 infection among prospectively recruited participants (N=1106). (A) Proportion of participants with fatigue (measured on FAS), dyspnea grade ≥1 (on mMRC), depression, anxiety, and stress (on DASS-21) over time, stratified by recovery status. BL: baseline (pre-COVID-19), W2: 2 weeks after infection, M1: 1 month, M3: 3 months, M6: 6 months, M9: 9 months, M12: 12 months. (B) Proportion of participants with problems in the five EQ-5D-5L domains over time, stratified by recovery status.

Based on exploratory multivariable regression analyses, we found evidence that fatigue (FAS) and pain or discomfort (EQ-5D-5L) prior to COVID-19, hospitalization due to COVID-19, and experiencing post-exertional malaise, chest pain, palpitations, arthralgia, cough, reduced concentration or memory, or visual disturbances at 6 months were associated with persistent non-recovery at 12 months (Figure 4, Supplementary Figure 8).

**Figure 4.**
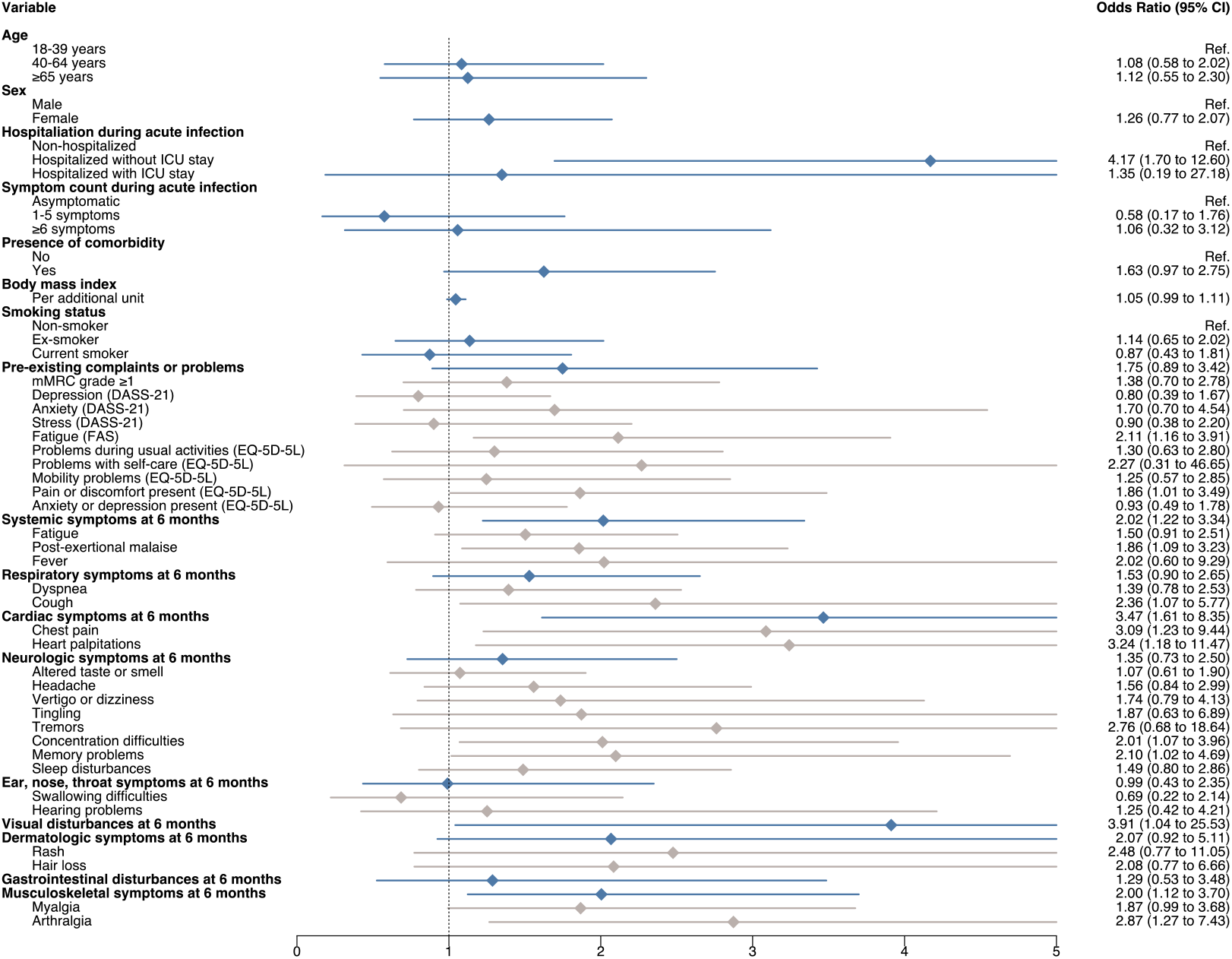
Associations for persistent non-recovery at 12 months after SARS-CoV-2 infection based on multivariable logistic regression models. Separate models were ran for each predictor variable with adjustment for age group, sex, hospitalization during acute COVID-19, symptom count and presence of comorbidities. Data from both retrospectively and prospectively recruited participants of the Zurich SARS-CoV-2 Cohort (N=1543) were used in all association analyses, with the exception of those relating to pre-existing complaints or health problems (data only available for prospectively recruited participants; N=1106). Blue lines represent estimates for grouped symptoms or complaints, while grey lines represent estimates for individual symptoms or complaints. OR: odds ratio; 95% CI: 95% confidence interval.

Among retrospectively recruited participants who reported non-recovery at 6 months and completed the 18-month questionnaire, 42% (37/88) had recovered by 18 months with an improvement in the overall health status in most participants (62%). However, COVID-19 related symptoms (52%) and other adverse health outcomes persisted in a large proportion of participants (Supplementary Figure 9).

## Discussion

To address the current knowledge gap regarding the natural course of post COVID-19 condition among affected individuals (11), we evaluated the longer-term recovery and health trajectories in a prospective, population-based cohort.

Consistent with previous studies, we found that 25% of previously unvaccinated individuals did not return to their normal health status by 6 months (1–3,25,4–7,10). Thereof, 37% fully recovered by 12 months and 42% by 18 months after infection. While we observed an initial worsening in participants’ HRQoL and health outcomes after infection, the proportion of participants reporting adverse health outcomes decreased over time. Individuals who did not require hospitalization during acute COVID-19 and individuals without pre-existing fatigue or pain or discomfort were more likely to experience full recovery within 12 months. Furthermore, specific systemic, cardiovascular, musculoskeletal symptoms at 6 months were also associated with non-recovery over this timeframe. Our findings imply that an important number of people may be affected by post COVID-19 condition and experience protracted health issues beyond one year. Meanwhile, the rates of recovery and the overall improvement in the severity of participants’ health impairment over time may also provide some hope for affected individuals. Identifying patient subgroups with the greatest risks of experiencing long-term health impairments and gaining a deeper understanding of the underlying mechanisms will be necessary for developing interventions targeting post COVID-19 condition.

Overall, approximately half of participants reported experiencing at least one symptom associated with post COVID-19 condition at 6 and 12 months after infection. However, symptom prevalence was relevantly lower with 27% and 25%, respectively, when restricting to symptoms that participants reported to be related to COVID-19. The comparative analysis leveraging data from a representative population-based seroprevalence study provided further evidence that overall symptom prevalence attributable to SARS-CoV-2 infection was up to 20%. The estimated excess risk of individual symptoms were relatively consistent with the proportion of symptoms self-reported by participants to be related to COVID-19. Yet, for several symptoms, we found insufficient evidence that their prevalence among infected individuals was elevated compared to non-infected individuals from the general population. Furthermore, some participants with symptoms after 12 months nevertheless reported full recovery, implying that the experienced symptoms may have only minimal impact on their daily lives. Considering that many published studies did not consider the relation of symptoms to COVID-19 or did not have a control group to estimate the proportion of adverse outcomes attributable to infection, findings from this study suggest that the reported estimates for post COVID-19 condition based on symptoms alone may be overestimated in the literature.

In this cohort, we used standardized assessment scales in addition to health status and self-reported symptoms, allowing to further contextualize our findings. The use of such measures increases comparability across studies and ensures that mental health aspects are also appropriately included. Based on DASS-21 and EQ-5D-5L, we found that the excess risk of experiencing symptoms of anxiety or depression among infected individuals was approximately 5-12%. This adds to the existing evidence of a relevant mental health burden of post COVID-19 condition (26,27), even in light of the generally increased prevalence of mental health-related issues in the general population during the pandemic (28). Given the size of the infected population globally, there is a need for healthcare services and policies to assess and address physical and mental health among those affected.

Altogether, our findings underscore the complexity in estimating the prevalence and trajectories of post COVID-19 condition and emphasize the importance of assessing multiple outcomes, using standardized scales, and including self-reported measures of recovery and health status for a comprehensive and person-centered perspective in clinical trials.

### Implications for trial design and outcome selection

Several clinical trials assessing potential interventions for post COVID-19 condition are currently underway (29). Evidence from this and other observational studies on its natural course can help to effectively design such trials. First, findings of this study provide important information on expected patient profiles and subgroups at greater risk of non-recovery. This may help in defining study populations in trials to ensure that individuals most likely to benefit from the intervention are included. Second, estimated rates of spontaneous recovery could be used to calculate necessary sample sizes to detect an improvement in participants’ outcomes. Third, findings from prospective cohorts may help to determine the most relevant outcome measures to assess post COVID-19 condition in clinical trials. Due to limited knowledge and emerging evidence, defining outcomes in this context has been challenging, leading to substantial heterogeneity across studies. Recently, a core outcome set for assessing post COVID-19 condition was suggested, informed by frequently reported symptoms in the literature and a multi-stakeholder modified Delphi process (17). The outcome measures used in our study are consistent with these recommendations. Meanwhile, our findings also emphasize the importance of using a patient-reported standardized measure of health status as a complementary outcome to capture the multidimensional impact of post COVID-19 condition on HRQoL and daily life functioning among affected individuals. Using such measures may also help to determine the overall benefit-harm balance of a potential intervention, especially in the presence of small effect sizes. Finally, evidence on the different symptoms experienced by individuals affected by post COVID-19 condition may help researchers to ensure that these are accurately and consistently captured in both arms of clinical trials at baseline and prospectively. This ensures that potential adverse effects of the intervention are appropriately investigated in the context of the underlying risk due to post COVID-19 condition.

### Limitations

Several limitations need to be considered when interpreting this study. First, we relied on the relative health status of participants to evaluate persistent health impairments after infection. We cannot exclude that the health status at the time of reporting may have reflected other infections or conditions. Second, our study relied solely on self-reported assessments. While we believe that standardized patient-reported outcome measures adequately reflect affected individuals’ experiences, future work should consider incorporating objective assessments and examining their correlation with self-reported measures. Third, we used pre-defined symptom lists to assess symptoms related to post COVID-19 condition. Although they were informed by what was most commonly reported by study participants and in the literature, these lists may represent only part of the symptoms experienced by those affected (30,31). We further did not assess their patterns, in particular whether they are relapsing, which has been shown to be a feature of post COVID-19 condition (32,33). Fourth, Zurich SARS-CoV-2 Cohort participants were enrolled when wildtype SARS-CoV-2 was the predominant circulating strain and prior to vaccination. Further research is needed to assess the effect of vaccination (34,35) or infection with emerging strains (e.g., Delta and Omicron) (36) on the development and persistence of post COVID-19 condition. Fifth, selection bias may have occurred if individuals who were more concerned with their health were more likely to participate, or if individuals with post COVID-19 condition were more likely to be retained in the study. This may have led to an overestimation of the prevalence of post COVID-19 condition. However, the high retention with minimal losses to follow-up strengthens our evaluations. Finally, we did not have information on whether participants were undergoing therapy for post COVID-19 condition during the study period. Thus, the natural course may not have been adequately reflected. However, given the current lack of effective evidence-based interventions, we deem it unlikely that our findings were affected by any such treatment.

## Conclusion

This population-based study demonstrated that despite a decrease in the severity of symptoms and health impairment over time, only two-fifths of those with post COVID-19 condition 6 months after infection fully recover within 18 months. Persisting health issues create significant challenges for affected individuals and their families and pose an important burden on population health and healthcare services. This underscores the value of effective infection prevention strategies and emphasizes the need for clinical trials aiming to establish effective interventions for post COVID-19 condition. Together with evidence from other observational studies, our findings demonstrate the importance of using multiple outcome measures and of considering the expected rates of natural recovery and heterogenous patients trajectories in the design and interpretation of future clinical trials.

## Data Availability

All data produced in the present study are available upon reasonable request to the authors.

## Author Contributions

Conception and design: T. Ballouz, D. Menges, J.S. Fehr, M.A. Puhan

Analysis and interpretation of the data: T. Ballouz, D. Menges, J.S. Fehr, M.A. Puhan

Drafting of the article: T. Ballouz

Critical revision for important intellectual content: T. Ballouz, D. Menges, A. Anagnostopoulos, A. Domenghino, H.E. Aschmann, A. Frei, J.S. Fehr, M.A. Puhan

Final approval of the article: T. Ballouz, D. Menges, A. Anagnostopoulos, A. Domenghino, H.E. Aschmann, A. Frei, J.S. Fehr, M.A. Puhan

Statistical expertise: T. Ballouz, D. Menges

Administrative, technical, or logistic support: T. Ballouz, D. Menges, A. Anagnostopoulos, A. Domenghino, H.E. Aschmann, A. Frei, J.S. Fehr, M.A. Puhan

Collection and assembly of data: T. Ballouz, D. Menges

## Notes

### Competing Interest Statement

The authors have declared no competing interest.

### Author Declarations

The study was approved by the ethics committee of the Canton of Zurich (BASEC 2020-01739).

