## Supplementary Material for "Natural course of post COVID-19 condition and implications for trial design and outcome selection: A population-based longitudinal cohort study"

### Supplementary Methods

#### Study design and participant recruitment

This analysis is based on the Zurich SARS-CoV-2 Cohort study, an ongoing, observational, population-based cohort study of individuals in the Canton of Zurich with polymerase chain reaction (PCR)-confirmed SARS-CoV-2 infection. We recruited participants through the Department of Health of the Canton of Zurich, which is notified of all diagnosed SARS-CoV-2 cases in the Canton through mandatory reporting. We evaluated all individuals for whom contact information was available for eligibility. Individuals who were 18 years or older, residing in the Canton of Zurich, able to follow study procedures, and having sufficient knowledge of the German language were eligible for participation. We obtained written or electronic consent from all participants. The study protocol was prospectively registered (ISRCTN14990068) and the study was approved by the ethics committee of the Canton of Zurich (BASEC 2020-01739).

#### Study population

We enrolled two populations of SARS-CoV-2 infected individuals. The first, referred to as *retrospectively recruited*, included all eligible individuals who were diagnosed with a SARS-CoV-2 infection prior to the start of the study (i.e., between 27 February 2020 and 05 August 2020). These individuals were enrolled between 06 October 2020 and 26 January 2021, at a median of 7.2 months after diagnosis of infection. The second, referred to as *prospectively recruited*, included an age-stratified (18 to 39 years, 40 to 64 years, 65+ years) random sample of all eligible individuals diagnosed between 06 August 2020 and 19 January 2021. These individuals were enrolled upon or shortly after diagnosis. Of note, we enrolled participants prior to the roll-out of COVID-19 vaccines and the emergence of SARS-CoV-2 variants of concern in Switzerland. Hence, all individuals were unvaccinated and had not been previously infected at the time of enrolment.

#### Control group population

To compare health outcomes among SARS-CoV-2 infected individuals with those in a comparable uninfected control population, we leveraged data from phase IV of the Corona Immunitas seroprevalence study ((1,2); study registration: ISRCTN18181860; ethics approval: BASEC 2020-01247). For this study, a total of 2'974 individuals were randomly selected from the general population of the Canton of Zurich using an age-stratified sampling strategy (20 to 64 years, 65+ years) and invited to participate. 697 agreed to take part (participation rate 23%) and were enrolled between 01 June and 12 July 2021. We excluded 66 participants who were likely to have been infected in the past or at the time of enrolment defined as: 1) self-reported history of a positive SARS-CoV-2 PCR or rapid antigen test, 2) positive anti-Spike (S) IgG or IgA antibody test with no history of vaccination, or 3) a positive anti-Nucleocapsid (N) IgG antibody test on enrolment. We also excluded 3 individuals who never completed the questionnaire, thereby including data from 628 non-infected individuals in this analysis.

#### Data sources

In the Zurich SARS-CoV-2 Cohort study, participants completed an electronic baseline questionnaire upon enrolment, which included questions on socio-demographics, pre-existing medical conditions, presence and severity of symptoms and hospitalizations during the acute infection, and health status prior to SARS-CoV-2 infection. Participants were asked to complete repeated follow-up questionnaires that included questions relating to their physical and mental health. Prospectively recruited answered the follow-up questionnaires at 2 weeks, 1, 3, 6, 9, and 12 months after infection while retrospectively recruited participants completed follow-up questionnaires at 9 and 12 months after infection. The 6-month and 12-month questionnaires were completed by 96% (1490/1543) and 91% (1407/1543) of the participants, respectively. Participants initially consented to participation for 12 months and were asked to consent for further biannual assessments upon completion of the 12-month assessment. At the time of writing, complete 18-month data was available only for the retrospectively recruited sample (318/411; 77% agreed to study prolongation and filled the questionnaire). In order to provide an even longer perspective of the progression of post COVID-19 condition, we additionally report outcomes at 18 months in the retrospectively recruited sample. Details on assessment timepoints and corresponding data for the two study populations are reported below in Supplementary Table 1.

The questionnaire of the phase IV Corona Immunitas seroprevalence study was closely aligned with the Zurich SARS-CoV-2 Cohort questionnaires to ensure comparability. It included identical questions on socio-demographics, pre-existing medical conditions, self-reported symptoms, and other health assessments as outlined in the section below. The timeframe of data collection (May to August 2021) corresponded to the timeframe of the 6-month and 12-month assessments of the prospectively and retrospectively recruited samples in the Zurich SARS-CoV-2 cohort, respectively. This was to minimize the influence of the timing of assessments on measured health outcomes (e.g., the influence of public health measures or duration of the pandemic on mental health outcomes).

We collected and managed all study data using the Research Electronic Data Capture (REDCap) system (3,4).

**Supplementary Table 1**. Data collected at each of the timepoints for the prospectively and retrospectively recruited populations.

| Follow-up | BL | W2 | M1 | M3 | M6 | M9 | M12 | M18 |
|---|---|---|---|---|---|---|---|---|
| **Symptoms** | Prosp. | Prosp. | Prosp. | Prosp. | Prosp. & retro. | Prosp. & retro. | Prosp. & retro. | Retro. |
| **FAS** | Prosp. (pre-COVID-19) | Prosp. | Prosp. | Prosp. | Prosp. & retro. | Prosp. & retro. | Prosp. & retro. | Retro. |
| **mMRC** | Prosp. (pre-COVID-19) | Prosp. | Prosp. | Prosp. | Prosp. & retro. | Prosp. & retro. | Prosp. & retro. | Retro. |
| **DASS-21** | Prosp. (pre-COVID-19) | Prosp. | Prosp. | Prosp. | Prosp. & retro. | Prosp. & retro. | Prosp. & retro. | Retro. |
| **EQ-5D-5L** | Prosp. (pre-COVID-19) | Prosp. | Prosp. | Prosp. | Prosp. & retro. | Prosp. & retro. | Prosp. & retro. | Retro. |

| | | | | | | | | |
|---|---|---|---|---|---|---|---|---|
| **EQ-VAS** | Prosp. (pre-COVID-19) | Prosp. | Prosp. | Prosp. | Prosp. & retro. | Prosp. & retro. | Prosp. & retro. | Retro. |

BL: baseline, W2: 2 weeks, M1: 1 month, M3: 3 months, M6: 6 months, M9: 9 months, M12: 12 months, M18: 18 months, Prosp.: prospectively recruited, Retro.: retrospectively recruited, FAS: Fatigue Assessment scale, mMRC: modified Medical Research Council (mMRC) dyspnea scale, DASS-21: 21-item Depression, Anxiety and Stress Scale, EQ-5D-5L: EuroQol 5-dimension 5-level, EQ-VAS: EuroQol Visual Analog Scale

#### Main outcomes

In the absence of an established core outcome set for post COVID-19 condition, we determined the outcomes based on several patient-relevant health measures and commonly reported symptoms (5,6) (Supplementary Table 2).

Our primary outcome was the overall relative health status of participants at 6 and 12 months after infection, defined using a combination of two self-reported measurements: 1) *(non)-recovery status* compared to usual health status before infection, and 2) *overall health* using the EuroQol visual analogue scale (EQ-VAS), with cut-off values for different levels of health impairment determined based on population normative values and studies on chronic-obstructive pulmonary disease (COPD) (7–9).

Secondary outcomes were the following:

1. Change in relative health status between 6 and 12 months after SARS-CoV-2 infection
2. Presence and severity of symptoms experienced within the past week using a comprehensive list of 23 predefined symptoms. We additionally asked participants to indicate whether they perceived the symptom to be related to COVID-19 to differentiate post COVID-19 symptoms from those due to other reasons.
3. Presence of fatigue, evaluated through the Fatigue Assessment Scale (FAS)
4. Presence and degree of functional disability due to dyspnea using the modified Medical Research Council (mMRC) dyspnea scale
5. Presence and severity of depression, anxiety and stress as determined by the 21-item Depression, Anxiety and Stress Scale (DASS-21)

6. Health-related quality of life (HRQoL) measured with the EuroQol 5-dimension 5-level (EQ-5D-5L) instrument.

Calculation of scores for assessment scales followed official guidance, as described previously (10–17).

**Supplementary Table 2.** Details on outcome measurement and definitions.

<table>
<tr><th>Outcome</th><th>Instrument</th><th>Definition/Categories</th></tr>
<tr><td colspan="3">Primary outcome</td></tr>
<tr><td>Overall relative health status</td><td>• Self-reported recovery status: “How do you feel now compared to when you were infected?” (fully recovered and symptom free, better but not fully recovered, neither better nor worse, worse)<br>• EQ-VAS</td><td>1. Recovered: participants reporting to be back to their normal health status<br>2. Non-recovered with mild health impairment: participants reporting not to be back to their normal health status and EQ-VAS scores >70<br>3. Non-recovered with moderate health impairment: EQ-VAS scores between 51–70<br>4. Non-recovered with severe health impairment: EQ-VAS scores of 50 or less</td></tr>
<tr><td colspan="3">Secondary outcomes</td></tr>
<tr><td>Change in relative health status</td><td>• Self-reported overall health status<br>• EQ-VAS</td><td>1. Recovered: participants reporting to be back to their normal health status<br>2. Improved: improvement in the health status category or in EQ-VAS by at least 8 points*<br>3. Worsened: deterioration of health status category or decrease in EQ-VAS by at least 8 points*<br>4. No change: no change in health category and changes in EQ-VAS of less than 8 points*</td></tr>
<tr><td>Symptoms</td><td>Self-reported symptoms:<br>«In the past 7 days, have you had one or more of the following symptoms that are unrelated to a chronic illness or allergy?»<br><br>“How bad or bothersome was this symptom for you? (Very severe, severe, moderate, mild, not bad at all)”<br><br>“Do you think this symptom is related to or a complication of coronavirus disease "COVID-19"? (No, Yes, I don’t know)”</td><td>• Presence of each symptom<br>• Severity of each symptom†<br>• Relation to COVID-19 for each symptom†</td></tr>
<tr><td>Fatigue</td><td>FAS</td><td>Presence of fatigue (defined as score ≥22)</td></tr>
</table>

| | | |
|---|---|---|
| Degree of functional disability due to dyspnea | mMRC dyspnea scale | Presence of dyspnea (defined as grade ≥1) |
| Depression, anxiety or stress | DASS-21 | Presence of depression (defined as score ≥10), anxiety (score ≥8), or stress (score ≥15) |
| Health-related quality of life (HRQoL) | EQ-5D-5L | Presence of problems in 5 dimensions (mobility, self-care, usual activities, depression or anxiety, pain or discomfort) |

*corresponding to minimally important difference (MID), if there is no category change. †Collected as of the 6-month follow-up in the questionnaire for prospectively recruited and 12-month follow-up in the questionnaire for retrospectively recruited participants. EQ-VAS: EuroQol visual analog scale; FAS: fatigue assessment scale; mMRC: modified Medical Research Council Dyspnea Scale; DASS-21: 21-item depression, anxiety and stress scale

As more evidence on post COVID-19 condition became available with time, we updated the predefined symptom list to ensure that the most relevant symptoms experienced by patients were captured. The change involved adding and removing certain symptoms and was implemented prior to the 6-month and 12-month follow-up of the prospectively and retrospectively recruited individuals, respectively.

- List of symptoms until the 3-month and 9-month follow-ups of the prospectively and retrospectively recruited individuals, respectively: fatigue, cough, dyspnea/shortness of breath, chest pain, nasal congestion, sore throat, sneezing, swollen lymph nodes, irritation of the eyes, abdominal pain, diarrhea, loss of appetite, nausea or vomiting, headache, myalgia, arthralgia, fever, altered taste and/or smell, rash, and tingling sensation of upper or lower extremities.
- Updated list of symptoms as of the 6-month and 12-month follow-ups of the prospectively and retrospectively recruited individuals, respectively: fatigue, post-exertional malaise, cough, dyspnea/shortness of breath, chest pain, heart palpitations, headache, myalgia, arthralgia, fever, gastrointestinal disturbances, altered taste and/or smell, difficulties swallowing, tingling sensation of upper or lower extremities, tremor, hearing problem, vertigo or dizziness, visual disturbances, rash, hair loss, memory problems, concentration difficulties, and sleep disturbances.

To improve comparability in estimates between prospectively and retrospectively recruited individuals, we used information from free text fields and data on symptom duration reported at the 12-month follow-up by the retrospectively recruited individuals to complete the 6-month and 9-month follow-up data (e.g., if symptoms were reported to have lasted for the past twelve months, the symptom was assumed to have been present also at earlier timepoints).

In addition, we grouped symptoms according to the body organ system involved as follows:

1. Systemic: fatigue, post-exertional malaise, fever
2. Respiratory: cough, dyspnea or shortness of breath
3. Cardiovascular: chest pain, heart palpitations
4. Neurologic: altered taste and/or smell, headache, concentration difficulties, memory problems, vertigo or dizziness, tremors, tingling sensation of extremities, sleep disturbances
5. Musculoskeletal: myalgia, arthralgia
6. Gastrointestinal: gastrointestinal disturbances
7. Ear, nose and throat: swallowing difficulties, hearing problems
8. Eye: visual disturbances
9. Dermatologic: skin rash, hair loss

For study participants reporting a reinfection over the study timeframe (N=27 by the 12-month follow-up and N=2 additional by the 18-month follow-up in retrospectively recruited individuals), defined as a new positive SARS-CoV-2 test more than 60 days after the initial positive test (18), we did not include data collected after the date of reinfection in our analysis to ensure that reported symptoms and health status are not related to potential new infections.

#### Statistical analysis

In line with our objectives, we performed three main analyses.

First, we descriptively analyzed the overall relative health status among infected individuals (all Zurich SARS-CoV-2 Cohort participants) at the 6-month follow-up and determined the proportion with complete recovery or mild, moderate, and severe health impairment. We performed multivariable regression analyses to assess the influence of potential predictors on non-recovery by 6 months. We included age group, sex, hospitalization, symptom count during the acute phase, and presence of comorbidities as a priori covariables. We based model selection on clinical reasoning and the Bayesian Information Criterion (BIC) by including variables that were reported to be associated with post COVID-19 condition and improved model fit by at least 2 points. We report regression analysis results as odds ratios (OR) with corresponding 95% confidence interval (CI). In addition, we calculated the prevalence and 95% Wilson CIs for each of the pre-defined symptoms at 6 and 12 months for the overall sample of infected individuals (Zurich SARS-CoV-2 Cohort), grouped based on the body organ system affected, as previously described.

Second, we estimated the excess risk of symptoms and adverse health outcomes in the SARS-CoV-2 infected population (Zurich SARS-CoV-2 Cohort) at the 6- and 12-month follow-ups compared to the general population (Corona Immunitas Zurich). We calculated adjusted risk differences (aRDs) using logistic regression models and included age group, sex, body mass index, and presence of comorbidities as a priori variables. The 95% CIs for the estimated aRDs were estimated using a bootstrap approach (resampling 1000 times at random). Since the data collection timeframe of the Corona Immunitas Zurich study corresponded to different follow-up timepoints of the prospectively (6 months) and retrospectively recruited (12 months) SARS-CoV-2 infected individuals, we performed these comparative analyses separately for the two groups.

Third, we focused on longer-term outcome trajectories among SARS-CoV-2 infected individuals within the Zurich SARS-CoV-2 Cohort who reported non-recovery at the 6-month follow-up. We evaluated the change in relative health status at 12 months. We then descriptively analyzed the proportion of participants reporting symptoms at the 6- and 12-month follow-up, individually and grouped by body organ system, and evaluated their co-occurrence to identify the most frequent phenotypes. Further, we descriptively compared the proportion of participants with adverse health outcomes (fatigue on FAS, dyspnea on mMRC, depression, anxiety, and stress on DASS-21, and health-related problems on EQ-5D-5L) prior to COVID-19 and up to 12 months in the non-recovered group of participants to those reporting recovery by 6 months. To identify potential predictors of persistence of non-recovery at the 12-month follow-up, we used multivariable logistic regression models. Separate models were run for each predictor variable and all models were adjusted for age group, sex, hospitalization during acute COVID-19, symptom count and presence of comorbidities. Model selection was performed based on knowledge of variables known to be associated with post COVID-19 condition and which improved the model fit (BIC) by at least two points. Additional predictor variables were body mass index, smoking status, pre-existing health problems (based on EQ-5D-5L) or symptoms (based on FAS, mMRC, DASS-21) prior to COVID-19, and the presence of symptoms at the 6-month follow-up. While we used data from both retrospectively and prospectively recruited participants of the Zurich SARS-CoV-2 Cohort (N=1543) in these association analyses, association analyses related to pre-existing complaints or health problems relied only on data from prospectively recruited participants (N=1106), since the respective information was collected only in this group. To test the robustness of our findings for the remaining variables (i.e., robustness of the assumption that the associations in the prospectively and retrospectively recruited samples are similar), we performed a sensitivity analysis in which we only included data from prospectively recruited participants for all predictor variables.

Finally, we descriptively analyzed the longitudinal trajectories of the aforementioned health outcomes at 18 months among retrospectively recruited participants who reported non-recovery at the 6-month follow-up.

All analyses were performed using R version 4.0.3 (19,20).

### Supplementary Results

**Supplementary Table 3.** Characteristics of participants in the two population samples enrolled in the Zurich SARS-CoV-2 Cohort.

| | **Overall (N=1543)** | **Prospectively recruited** (diagnosed 06 Aug 2020- 19 Jan 2021) **(N=1106)** | **Retrospectively recruited** (diagnosed 27 Feb 2020- 05 Aug 2020) **(N=437)** |
|---|---|---|---|
| **Age (years),** Median (IQR) | 49.0 (34.0 to 63.0) | 50.0 (35.0 to 66.0) | 47.0 (33.0 to 58.0) |
| **Age group** | | | |
| 18-39 years | 510 (33.1%) | 344 (31.1%) | 166 (38.0%) |
| 40-64 years | 656 (42.5%) | 449 (40.6%) | 207 (47.4%) |
| 65+ years | 377 (24.4%) | 313 (28.3%) | 64 (14.6%) |
| **Female sex**, N (%) | 781 (50.6%) | 566 (51.2%) | 215 (49.2%) |
| **Initial symptom severity** | | | |
| Asymptomatic | 195 (12.8%) | 148 (13.5%) | 47 (10.9%) |
| Mild to moderate | 942 (61.8%) | 721 (66.0%) | 221 (51.2%) |
| Severe to very severe | 388 (25.4%) | 224 (20.5%) | 164 (38.0%) |
| Missing | 18 | 13 | 5 |
| **Initial symptom count,** Median (IQR) | 5.0 (3.0 to 8.0) | 5.0 (3.0 to 8.0) | 6.0 (3.0 to 8.0) |
| **Symptoms experienced during acute COVID-19** | | | |
| Fatigue | 935 (62.9%) | 661 (63.0%) | 274 (62.7%) |
| Fever | 860 (58.5%) | 589 (57.0%) | 271 (62.0%) |
| Headache | 806 (54.9%) | 606 (58.8%) | 200 (45.8%) |
| Cough | 754 (51.3%) | 540 (52.3%) | 214 (49.0%) |
| Myalgia | 701 (47.9%) | 495 (48.2%) | 206 (47.1%) |
| Altered taste or smell | 653 (44.8%) | 442 (43.3%) | 211 (48.3%) |
| Nasal congestion | 521 (36.1%) | 424 (42.1%) | 97 (22.2%) |
| Sore throat | 459 (32.0%) | 324 (32.5%) | 135 (30.9%) |
| Dyspnea or shortness of breath | 358 (25.0%) | 234 (23.5%) | 124 (28.4%) |
| Appetite loss | 385 (26.6%) | 259 (25.6%) | 126 (28.8%) |
| Sneezing | 295 (20.7%) | 245 (24.8%) | 50 (11.4%) |
| Arthralgia | 261 (18.3%) | 178 (18.0%) | 83 (19.0%) |
| Diarrhea | 249 (17.4%) | 181 (18.2%) | 68 (15.6%) |
| Chest pain | 223 (15.6%) | 157 (15.8%) | 66 (15.1%) |
| Irritated eyes | 180 (12.7%) | 138 (14.0%) | 42 (9.6%) |
| Nausea or vomiting | 161 (11.3%) | 123 (12.4%) | 38 (8.7%) |
| Abdominal pain | 98 (6.9%) | 71 (7.2%) | 27 (6.2%) |
| Tingling sensation of extremities | 90 (6.4%) | 63 (6.5%) | 27 (6.2%) |
| Swollen lymph nodes | 73 (5.2%) | 55 (5.6%) | 18 (4.1%) |
| Skin rash | 61 (4.3%) | 44 (4.5%) | 17 (3.9%) |
| **Hospitalization and ICU stay** | | | |
| Non-hospitalized | 1401 (91.5%) | 1051 (95.6%) | 350 (81.0%) |
| Hospitalized without ICU stay | 115 (7.5%) | 44 (4.0%) | 71 (16.4%) |
| Hospitalized with ICU stay | 15 (1.0%) | 4 (0.4%) | 11 (2.5%) |

| | | | |
|---|---|---|---|
| *Intubation during ICU stay* | 8 (53.3%) | 1 (25%) | 7 (63.6%) |
| Missing | 12 | 7 | 5 |
| **At least one medical comorbidity** | 471 (30.8%) | 324 (29.5%) | 147 (34.2%) |
| Missing | 16 | 9 | 7 |
| **Monthly household income** | | | |
| <6'000 CHF | 489 (33.8%) | 356 (34.2%) | 133 (32.8%) |
| 6'000 - 12'000 CHF | 614 (42.4%) | 458 (44.0%) | 156 (38.4%) |
| >12'000 CHF | 344 (23.8%) | 227 (21.8%) | 117 (28.8%) |
| Missing | 96 | 65 | 31 |
| **Current employment status** | | | |
| Employed | 1027 (67.7%) | 706 (64.7%) | 321 (75.4%) |
| Student | 67 (4.4%) | 53 (4.9%) | 14 (3.3%) |
| Retired | 327 (21.5%) | 265 (24.3%) | 62 (14.6%) |
| Unemployed or other | 97 (6.4%) | 68 (6.2%) | 29 (6.8%) |
| Missing | 25 | 14 | 11 |
| **Highest education level reached** | | | |
| None or mandatory school | 65 (4.3%) | 45 (4.1%) | 20 (4.7%) |
| Vocational training or specialized baccalaureate | 645 (42.5%) | 459 (42.1%) | 186 (43.5%) |
| Higher technical school or college | 395 (26.0%) | 289 (26.5%) | 106 (24.8%) |
| University | 412 (27.2%) | 296 (27.2%) | 116 (27.1%) |
| Missing | 26 | 17 | 9 |
| **Swiss nationality** | 1309 (84.8%) | 943 (85.3%) | 366 (83.3%) |
| **Smoking status** | | | |
| Non-smoker | 901 (59.3%) | 656 (60.1%) | 245 (57.2%) |
| Ex-smoker | 410 (27.0%) | 288 (26.4%) | 122 (28.5%) |
| Smoker | 209 (13.8%) | 148 (13.6%) | 61 (14.3%) |
| Missing | 23 | 14 | 10 |
| **Body mass index,** Median (IQR) | 24.4 (22.0 to 26.8) | 24.2 (21.9 to 26.6) | 24.8 (22.2 to 27.5) |

IQR: interquartile range, N: number, ICU: intensive care unit, CHF: swiss francs

**Supplementary Table 4.** Characteristics of participants recovering at 6 months compared to those reporting non-recovery in the Zurich SARS-CoV-2 Cohort (Total N= 1543).

| | **Recovered (N=1070)** | **Non-recovered (N=348)** | **Missing (N=125)** |
|---|---|---|---|
| **Age (years),** Median (IQR) | 48.0 (33.0 to 62.0) | 53.5 (41.8 to 68.0) | 48.0 (32.0 to 60.0) |
| **Age group** | | | |
| 18-39 years | 382 (35.7%) | 79 (22.7%) | 49 (39.2%) |
| 40-64 years | 451 (42.1%) | 159 (45.7%) | 46 (36.8%) |
| 65+ years | 237 (22.1%) | 110 (31.6%) | 30 (24.0%) |
| **Female sex**, N (%) | 513 (47.9%) | 203 (58.3%) | 65 (52.0%) |
| **At least one medical comorbidity** | 286 (26.8%) | 154 (44.3%) | 31 (28.2%) |
| Missing | 1 | 0 | 15 |
| **Hospitalization and ICU stay** | | | |
| Hospitalized | 64 (6.0%) | 54 (15.5%) | 12 (10.6%) |
| Non-hospitalized | 1006 (94.0%) | 294 (84.5%) | 101 (89.4%) |
| Missing | 0 (0%) | 0 (0%) | 12 |
| **Initial symptom count,** Median (IQR) | 5.0 (3.0 to 7.0) | 7.0 (5.0 to 10.0) | 0.0 (0.0 to 5.0) |

IQR: interquartile range, N: number, ICU: intensive care unit

**Supplementary Table 5.** Odds ratios for self-reported symptoms and adverse health outcomes in SARS-CoV-2 infected individuals at 6 months (N= 1106) and 12 months (N= 437; Zurich SARS-CoV-2 Cohort) compared with 628 non-infected individuals from the general population (Corona Immunitas Zurich). Results were calculated using logistic regression models adjusted for age group, sex, body mass index and presence of chronic comorbidities.

| | 6 months (prospectively recruited) | | 12 months (retrospectively recruited) | |
|---|---|---|---|---|
| **Outcome** | **OR (95% CI)** | **p-value** | **OR (95% CI)** | **p-value** |
| **Symptoms** | | | | |
| Taste or smell alterations | 21.20 (7.90 to 86.66) | <0.001 | 16.20 (5.25 to 71.50) | <0.001 |
| Swallowing difficulties | 4.55 (2.00 to 12.27) | 0.001 | 2.50 (0.81 to 8.40) | 0.119 |
| Chest pain | 4.46 (1.96 to 12.05) | 0.001 | 4.58 (1.74 to 13.74) | 0.003 |
| Hair loss | 4.28 (2.21 to 9.14) | <0.001 | 2.35 (0.99 to 5.89) | 0.059 |
| Concentration difficulties | 4.16 (2.48 to 7.45) | <0.001 | 3.74 (2.00 to 7.30) | <0.001 |
| Dyspnea or shortness of breath | 4.03 (2.45 to 6.95) | <0.001 | 2.88 (1.53 to 5.61) | 0.001 |
| Post-exertional malaise | 3.14 (2.16 to 4.65) | <0.001 | 2.64 (1.66 to 4.25) | <0.001 |
| Memory problems | 2.96 (1.77 to 5.22) | <0.001 | 4.31 (2.25 to 8.53) | <0.001 |
| Heart palpitations | 2.67 (1.48 to 5.13) | 0.002 | 3.30 (1.62 to 7.04) | 0.001 |
| Visual disturbances | 1.91 (0.97 to 3.97) | 0.070 | 1.63 (0.69 to 3.98) | 0.269 |
| Tremors | 1.82 (0.78 to 4.75) | 0.190 | 0.63 (0.15 to 2.38) | 0.507 |
| Vertigo or dizziness | 1.79 (1.14 to 2.89) | 0.014 | 2.32 (1.26 to 4.31) | 0.007 |
| Fatigue | 1.73 (1.28 to 2.37) | 0.001 | 1.71 (1.17 to 2.49) | 0.005 |
| Gastrointestinal disturbances | 1.44 (0.92 to 2.29) | 0.121 | 1.47 (0.77 to 2.79) | 0.237 |
| Skin rash | 1.43 (0.72 to 3.00) | 0.324 | 1.88 (0.82 to 4.47) | 0.140 |
| Sleep disturbances | 1.40 (0.99 to 1.99) | 0.058 | 1.66 (1.07 to 2.59) | 0.024 |
| Cough | 1.35 (0.84 to 2.24) | 0.226 | 1.79 (0.98 to 3.31) | 0.061 |
| Hearing problems | 1.14 (0.63 to 2.11) | 0.668 | 0.74 (0.28 to 1.78) | 0.510 |
| Tingling sensation of extremities | 1.14 (0.67 to 1.98) | 0.642 | 1.05 (0.51 to 2.14) | 0.893 |
| Myalgia | 1.06 (0.75 to 1.51) | 0.754 | 0.94 (0.58 to 1.51) | 0.800 |
| Headache | 0.98 (0.70 to 1.38) | 0.906 | 1.19 (0.79 to 1.79) | 0.415 |
| Arthralgia | 0.92 (0.60 to 1.43) | 0.716 | 1.41 (0.81 to 2.45) | 0.221 |
| Fever | 0.50 (0.19 to 1.32) | 0.156 | 1.04 (0.40 to 2.73) | 0.935 |
| **Adverse health outcomes** | | | | |
| Depression (DASS-21) | 1.03 (0.77 to 1.40) | 0.824 | 1.16 (0.79 to 1.69) | 0.446 |
| Anxiety (DASS-21) | 1.72 (1.19 to 2.52) | 0.004 | 2.53 (1.64 to 3.94) | <0.001 |
| Stress (DASS-21) | 1.08 (0.73 to 1.60) | 0.709 | 1.47 (0.93 to 2.32) | 0.100 |

| | | | | |
|---|---|---|---|---|
| Mobility problems (EQ-5D-5L) | 1.13 (0.81 to 1.59) | 0.477 | 1.56 (0.98 to 2.47) | 0.059 |
| Problems during usual activities (EQ-5D-5L) | 1.49 (1.03 to 2.19) | 0.037 | 2.04 (1.31 to 3.21) | 0.002 |
| Self-care problems (EQ-5D-5L) | 1.94 (0.79 to 5.22) | 0.161 | 1.85 (0.60 to 5.90) | 0.287 |
| Anxiety or depression (EQ-5D-5L) | 1.02 (0.78 to 1.32) | 0.898 | 0.82 (0.60 to 1.11) | 0.204 |
| Pain or discomfort (EQ-5D-5L) | 0.72 (0.58 to 0.91) | 0.006 | 1.35 (0.98 to 1.88) | 0.069 |
| Fatigue (FAS) | 0.80 (0.65 to 0.99) | 0.045 | 1.22 (0.92 to 1.61) | 0.166 |
| Extreme fatigue (FAS) | 1.10 (0.53 to 2.43) | 0.798 | 0.72 (0.25 to 2.01) | 0.541 |
| mMRC grade ≥1 (mMRC) | 1.14 (0.86 to 1.54) | 0.362 | 1.38 (0.94 to 2.04) | 0.104 |
| mMRC grade ≥2 (mMRC) | 0.95 (0.45 to 2.08) | 0.902 | 1.54 (0.59 to 3.96) | 0.374 |

OR: odds ratio, CI: confidence interval, DASS-21: 21-item Depression, Anxiety, and Stress Scale, EQ-5D-5L: EuroQol 5-dimension 5-level, FAS: Fatigue Assessment Scale, mMRC: modified Medical Research Council dyspnea scale

**Supplementary Figure 1.** Study enrolment into the Zurich SARS-CoV-2 Cohort.

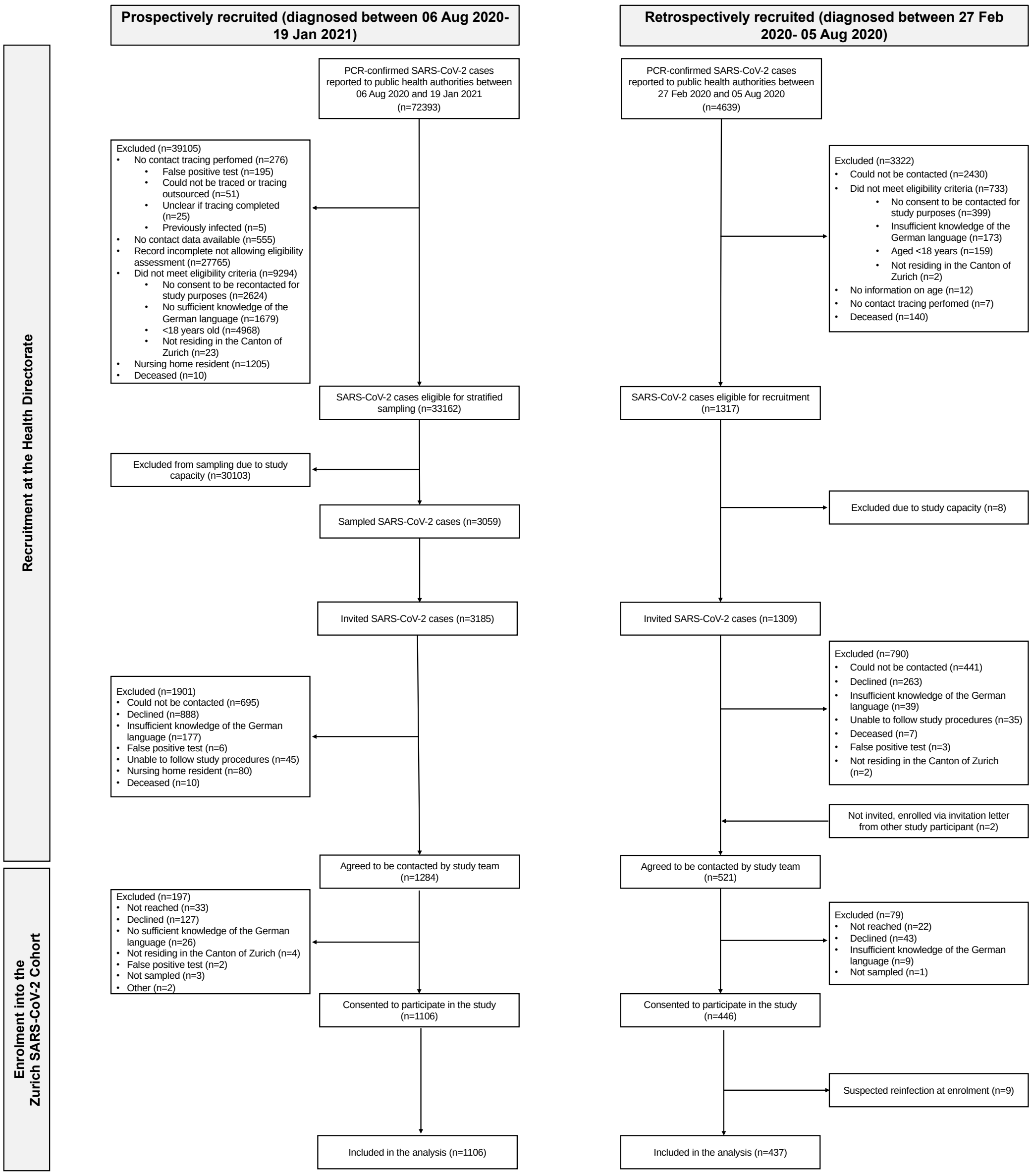


We included data from 1543 SARS-CoV-2 infected participants in this study. Among 4639 individuals diagnosed with SARS-CoV-2 infection between 27 February and 05 August 2020 (retrospectively recruited), 1309 were eligible and invited to participate in the study. 446 agreed to participate (participation rate 34.1%) and 437 were included in this analysis (9 excluded since reporting reinfection at enrolment). Among 72393 individuals diagnosed with SARS-CoV-2 infection between 06 August 2020 and 19 January 2021 (prospectively recruited), 3185 were eligible and invited. 1106 individuals agreed to participate and were included in this analysis (participation rate 34.7%).

A SARS-CoV-2 reinfection was reported by 13 participants at 6 months, by 3 participants at 9 months, by 11 participants at 12 months and by 2 additional participants in the retrospectively recruited sample at 18 months (total 29 participants). Data collected after the reinfection timepoint was omitted for these participants.

**Supplementary Figure 2.** Factors associated with non-recovery at 6 months in the Zurich SARS-CoV-2 Cohort (N=1543). Model adjusted for age, sex, hospitalization and symptom count during acute COVID-19 and presence of chronic comorbidity. According to the regression model, those at lowest risk (aged 18-39 years, male, had no comorbidities, and were asymptomatic and non-hospitalized during acute COVID-19) had a risk of 5.1% of being non-recovered whereas those at highest risk (aged 65+ years, female, with at least one comorbidity, were hospitalized during acute COVID-19 and had 6 symptoms or more) had a risk of 69%.

CI: confidence interval; Ref: reference.

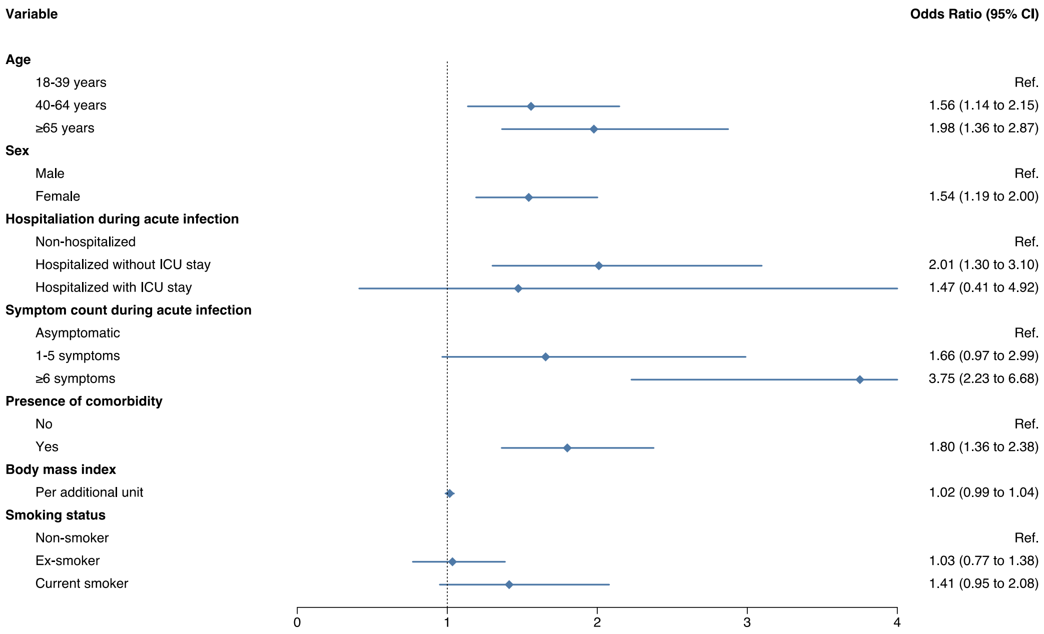

**Supplementary Figure 3.** Prevalence and severity of 23 prespecified symptoms at 6 and 12 months after SARS-CoV-2 infection, grouped by body organ system affected among the prospectively recruited cohort (N=1106). Error bars show the prevalence and 95% confidence interval for each of the symptoms. Solid line refers to symptoms reported by the participants to be related to COVID-19. Dotted line refers to symptoms regardless of their relation to COVID-19. Stacked bar plot shows self-reported severity of symptoms.

M6: 6 months after infection, M12: 12 months, Resp.: respiratory, Cardio.: cardiovascular, Neuro.: neurologic, Musculo.: musculoskeletal, GI: gastrointestinal, ENT: ear, nose and throat, Dermato: dermatologic, diff.: difficulties.

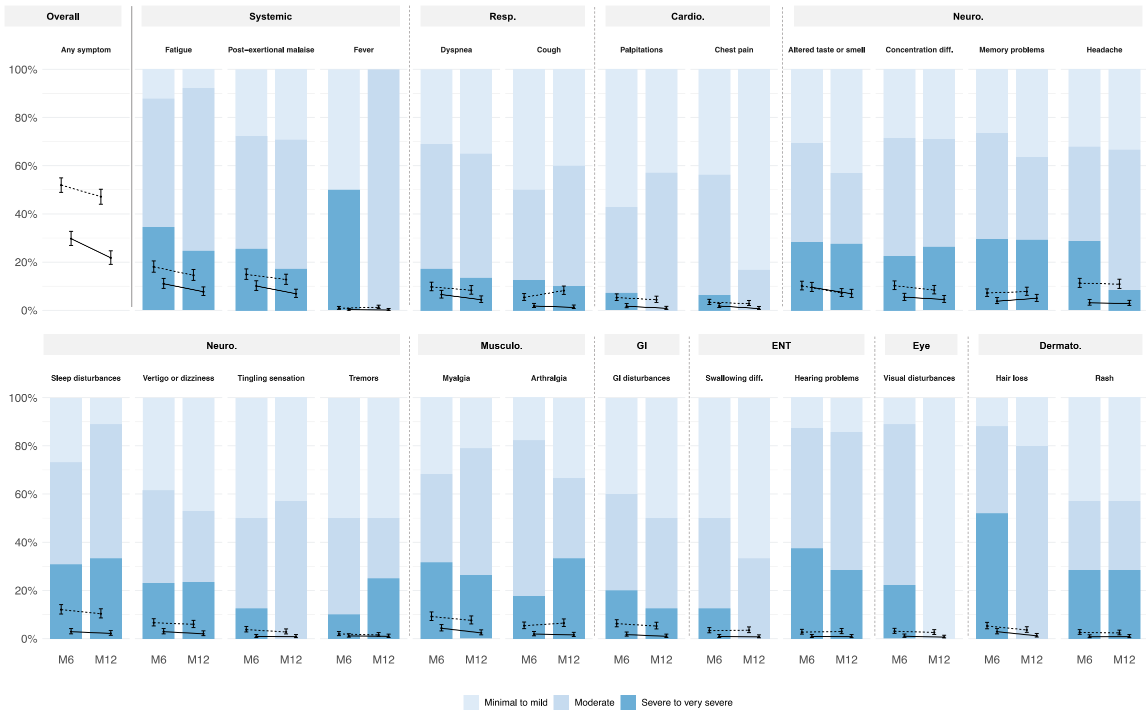

**Supplementary Figure 4.** (A) Prevalence and excess risk of adverse health outcomes using the scale-based assessments at 6 months in the prospectively recruited infected group (N=1106; Zurich SARS-CoV-2 Cohort) compared to the non-infected individuals from the general population (N=628; Corona Immunitas Zurich). (B) Prevalence and excess risk of adverse health outcomes using the scale-based assessments at 12 months in the retrospectively recruited infected group (N=437; Zurich SARS-CoV-2 Cohort) compared to non-infected individuals (N=628; Corona Immunitas Zurich).

DASS-21: 21-item Depression, Anxiety, and Stress Scale, EQ-5D-5L: EuroQol 5-dimension 5-level, FAS: Fatigue Assessment Scale, mMRC: modified Medical Research Council dyspnea scale.

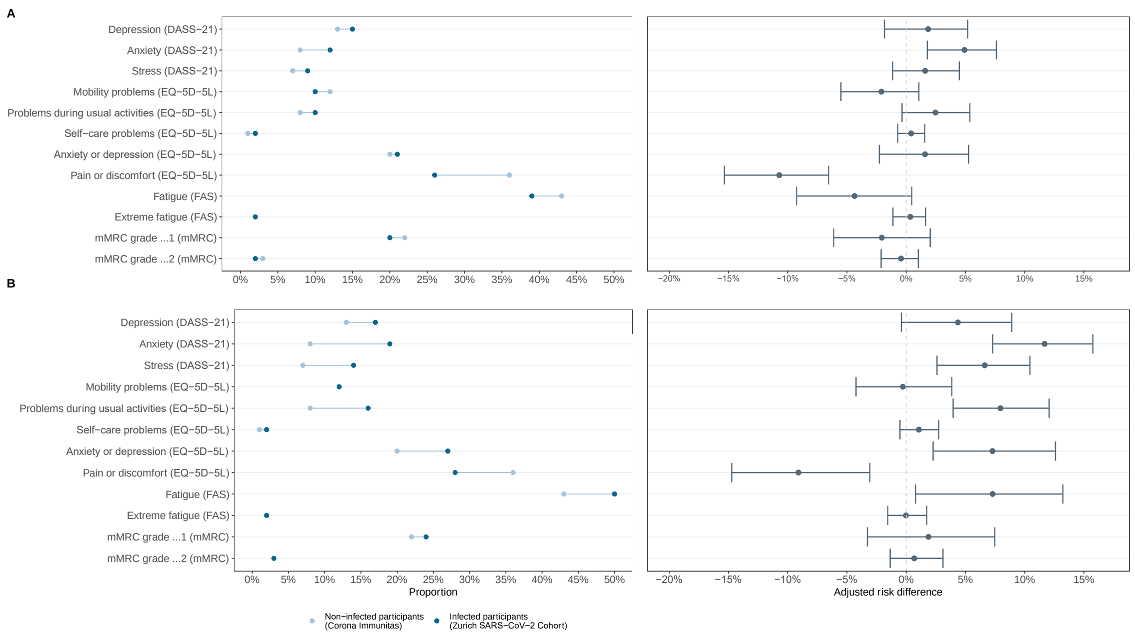

**Supplementary Figure 5**. Change in health status between the 6-month and the 12-month follow-up among participants who reported non-recovery at the 6-month follow-up, stratified by the severity of health impairment (N=348; Zurich SARS-CoV-2 Cohort).

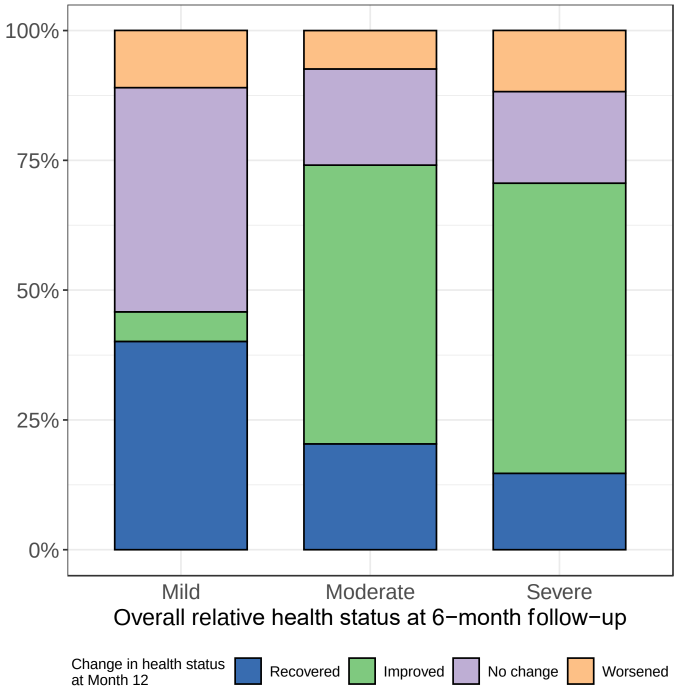

**Supplementary Figure 6**. Prevalence and severity of 23 prespecified symptoms at 6 and 12 months after SARS-CoV-2 infection among participants reporting non-recovery at the 6-month follow-up (N=348), grouped by body organ system affected. Error bars show the prevalence and 95% confidence interval for each of the symptoms among all non-recovered participants in the Zurich SARS-CoV-2 cohort. Solid line refers to symptoms reported by the participants to be related to COVID-19. Dotted line refers to symptoms regardless of their relation to COVID-19. Stacked bar plot shows self-reported severity of symptoms in prospectively recruited participants (N=238).

M6: 6 months after infection, M12: 12 months, Resp.: respiratory, Cardio.: cardiovascular, Neuro.: neurologic, Musculo.: musculoskeletal, GI: gastrointestinal, ENT: ear, nose and throat, Dermato: dermatologic, diff.: difficulties.

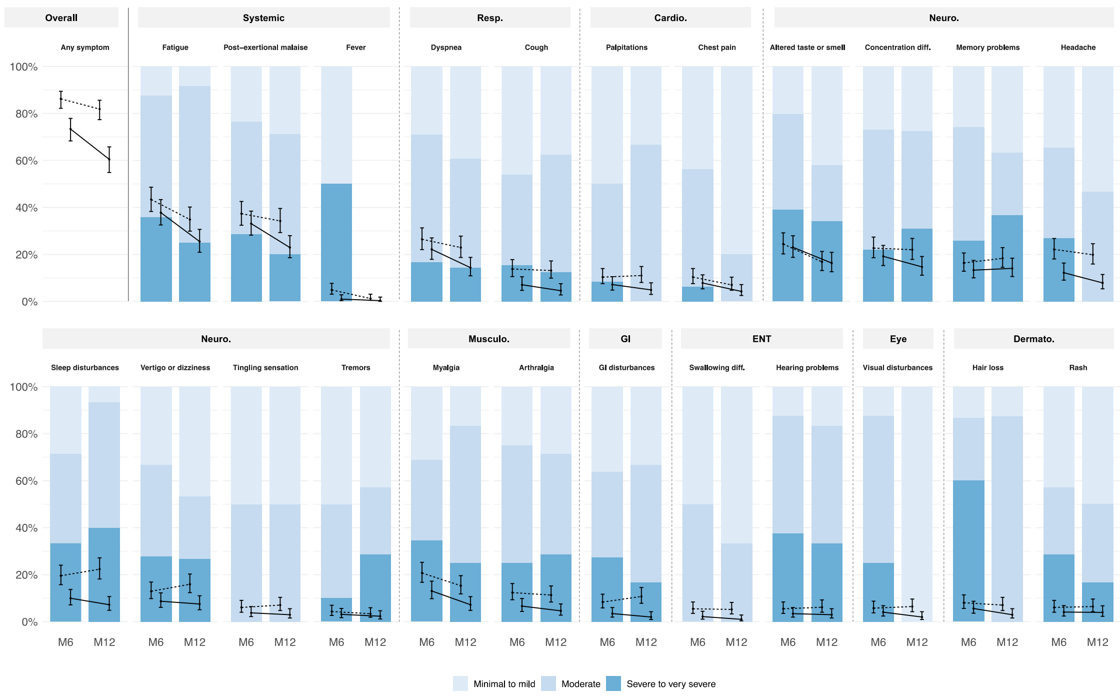

**Supplementary Figure 7.** Most common co-occurrence patterns of COVID-19 related symptoms grouped by body organ system at 6 months (panel A) and 12 months (panel B) after infection among SARS-CoV-2 infected individuals reporting non-recovery at the 6-month follow-up (N=348; Zurich SARS-CoV-2 Cohort). The presence of least one symptom in the corresponding body organ system is indicated by the blue dots. Black lines connecting the blue dots indicate co-occurrence, and bars show the number of individuals with the corresponding co-occurrence pattern of symptoms.

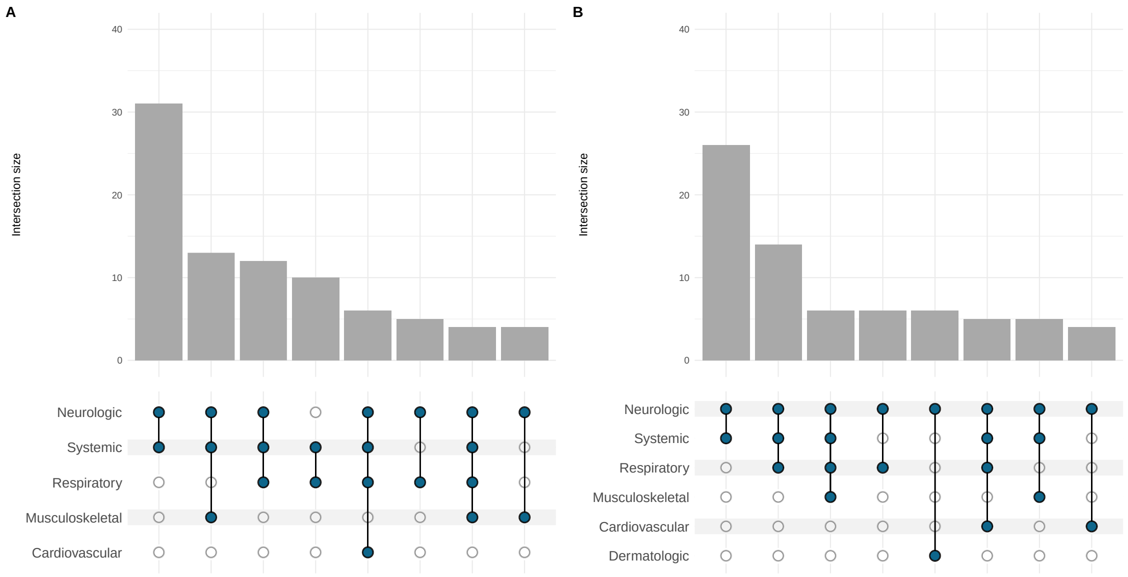

**Supplementary Figure 8.** Associations for persistent non-recovery among the prospectively recruited sample (N=1106) at 12 months after SARS-CoV-2 infection based on multivariable logistic regression models (sensitivity analysis). Separate models were run for each predictor variable and all models were adjusted for age group, sex, hospitalization during acute COVID-19, symptom count and presence of comorbidities. Blue lines represent estimates for grouped symptoms or complaints, while grey lines represent estimates for individual symptoms or complaints. OR: odds ratio; 95% CI: 95% confidence interval.

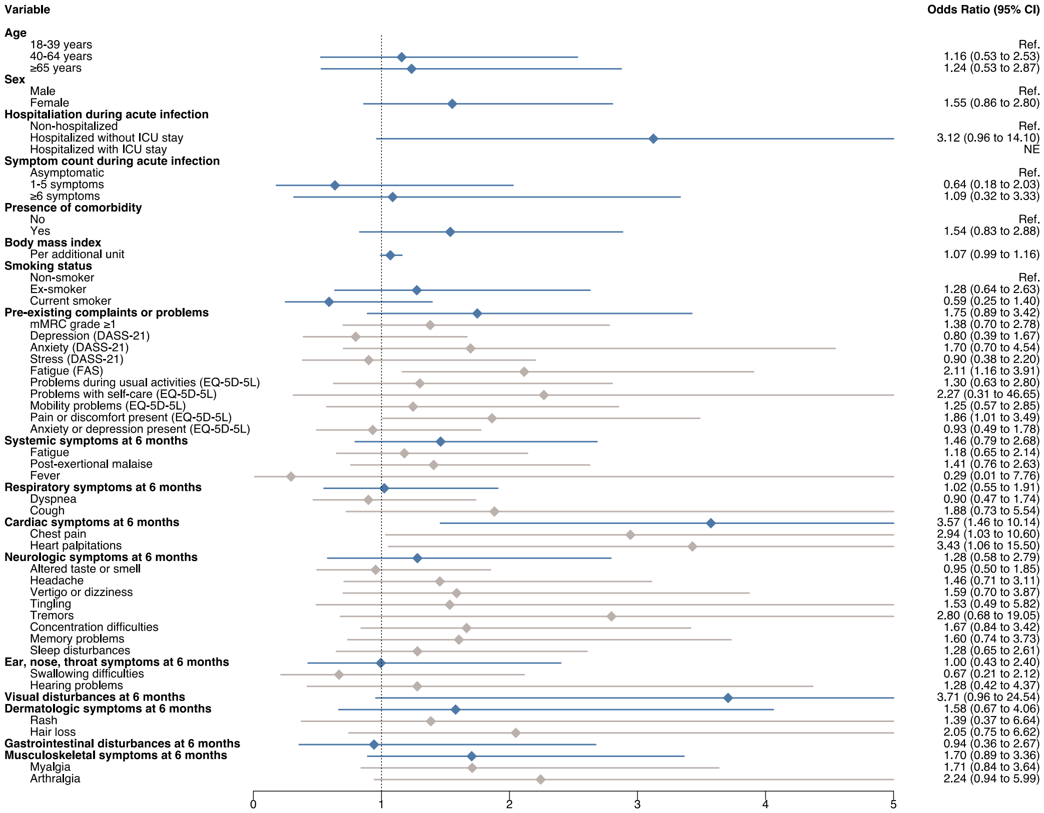

**Supplementary Figure 9.** Adverse health outcomes stratified by recovery status at 6 months after SARS-CoV-2 infection among retrospectively recruited participants who consented to study prolongation and completed the 18-month follow-up (N=318). (A) Proportion of participants with fatigue (measured on FAS), dyspnea grade ≥1 (on mMRC), depression, anxiety and stress (on DASS-21) over time, stratified by recovery status. (B) Proportion of participants with problems in the 5 EQ-5D-5L domains, stratified by recovery status.

M6: 6 months after infection, M12: 12 months, M18: 18 months.

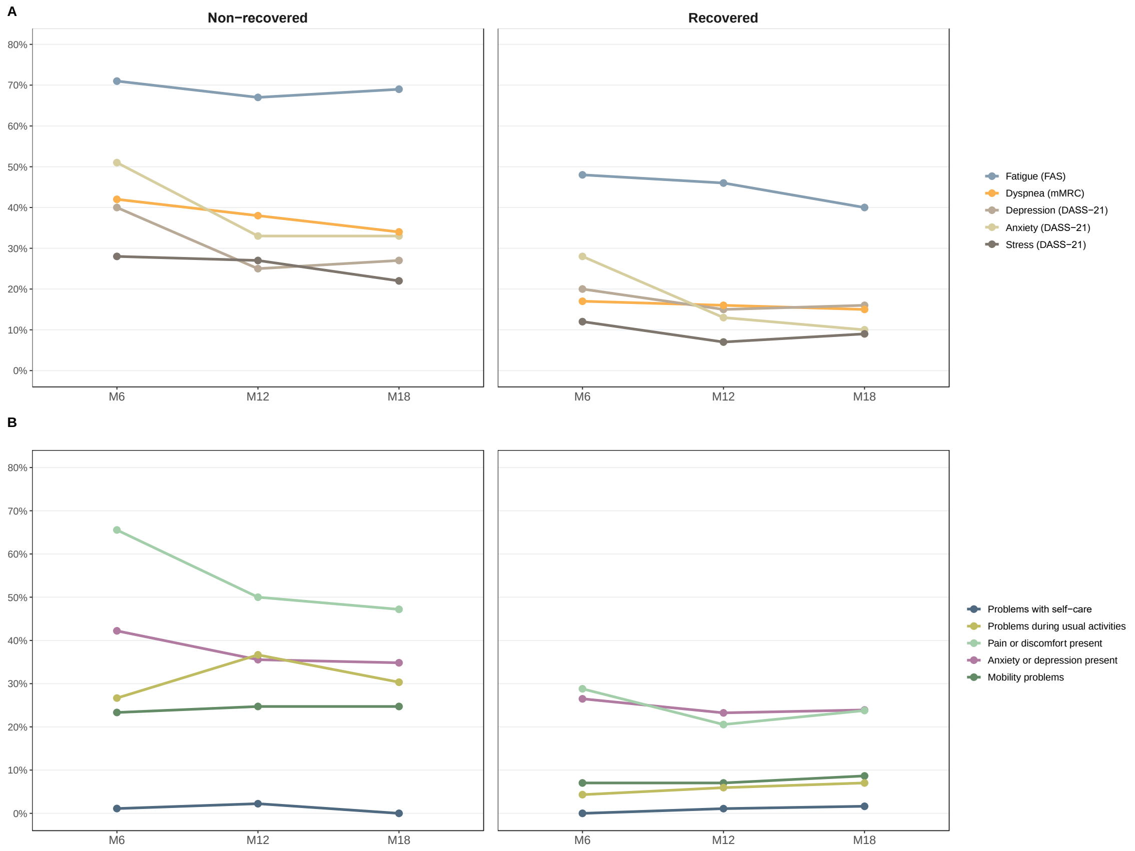

18. European Centre for Disease Prevention and Control. Reinfection with SARS-CoV-2: implementation of a surveillance case definition within the EU/EEA [Internet]. Available from: https://www.ecdc.europa.eu/en/publications-data/reinfection-sars-cov-2-implementation-surveillance-case-definition-within-eueea

19. R Core Team. R: a language and environment for statistical computing [Internet]. 2018 [cited 2021 Nov 19]. Available from: Http://www.r-project.org/

20. R Core Team. R: A language and environment for statistical computing. R Foundation for Statistical Computing [Internet]. Vienna, Austria. 2021. Available from: https://www. R-project.org/
